# Local ancestry-informed rare variant burden testing improves gene discovery in admixed populations

**DOI:** 10.64898/2026.07.13.26357993

**Authors:** Pragati Kore, Taotao Tan, Wenxuan Lu, Astrid Manuel-Friedman, Linfeng Hu, Nilanjan Chatterjee, Wei Zhou, Ryan S. Dhindsa, Elizabeth G. Atkinson

## Abstract

Rare-variant association studies enable the discovery of high-impact genetic contributors often missed by conventional genome-wide association studies focused on common variation. However, standard burden tests aggregate variants without accounting for local ancestry in admixed genomes, reducing power when rare variant frequencies or genetic effects differ across ancestral backgrounds. Here, we introduce Tractor-Burden, an ancestry-aware gene-based association method that partitions rare-variant burden by inferred local ancestry and estimates ancestry-specific effects within a unified regression. In simulations, Tractor-Burden is well calibrated and improves power over standard burden tests under effect heterogeneity. Applied to whole-genome sequencing data from 47,152 admixed African-European individuals in the All of Us Research Program, Tractor-Burden recapitulates known associations, including ancestry-enriched effects at *LDLR*, and identifies additional suggestive genes and pathways for type 2 diabetes. Tractor-Burden extends rare-variant association testing to admixed genomes and provides a scalable framework for detecting and interpreting gene-level effects across local ancestry backgrounds.

## Introduction

Over the past decades, advances in genomic technologies and the growth of large-scale biobanks have substantially improved our understanding of the genetic architecture underlying human disease and biological traits^1,2^. Common variant genome-wide association studies (GWAS) have identified thousands of loci associated with complex diseases, providing important insight into disease biology, therapeutic targeting, and polygenic risk prediction^3,4^. Although GWAS have successfully identified many disease-associated loci, much of the heritability underlying complex traits remains unexplained^3,5^. Increasingly, large-scale sequencing studies have shifted toward rare coding variation^6^, which can confer larger effect sizes and offer more direct functional interpretation than many common variant associations^7^. However, despite these advances, most large-scale genomic studies remain heavily biased toward populations of European ancestry^8,9^. Furthermore, most rare variant analytical frameworks remain designed for homogeneous populations, limiting the transferability of genetic findings across groups and contributing to disparities in genomic discovery and clinical interpretation^10,11^. As genomic datasets continue to diversify, understanding complex trait genetics across ancestrally diverse and admixed populations has become increasingly important for equitable precision medicine^12^.

Standard approaches to control for population structure in rare variant association studies (RVAS), including aggregate burden and kernel-based approaches such as CMC^13^, SKAT^14^, SKAT-O^15^, and mixed-model frameworks like SAIGE-GENE^16^ and SAIGE-GENE+^17^, typically rely on global ancestry estimates, such as principal components or admixture proportions. While these methods capture broad-scale variation, they do not account for local ancestry^18^, which reflects the ancestry of specific chromosomal segments within admixed genomes (Fig. 1). In admixed genomes, each chromosomal segment is assigned a local ancestry corresponding to the ancestral population from which it was inherited. Rare variants are often geographically or ancestry-enriched, and their frequencies, haplotype backgrounds, and effects may differ across local ancestry tracts^19–21^. Burden tests for rare variant analysis aggregate all qualifying variants into a single gene-level count and merge signals arising from distinct ancestry backgrounds, therefore reducing power when effects are heterogeneous and obscuring interpretation when an association is driven by ancestry-specific burden^22^. As a result, ancestry-enriched rare variant signals may become diluted when variants are collapsed uniformly across heterogeneous ancestral tracts.

**Figure 1.**
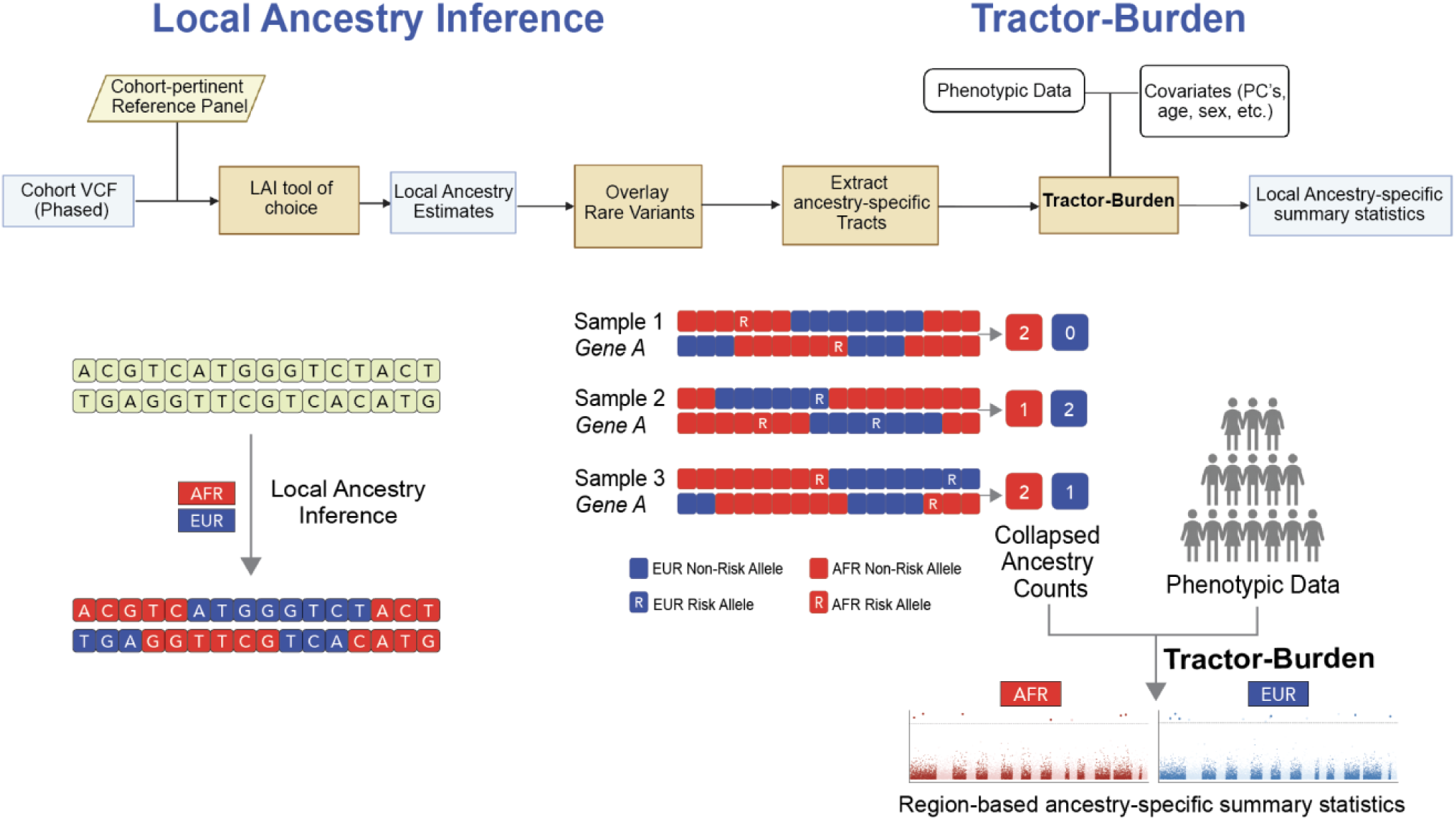
Overview of Tractor-Burden framework^32^. Local ancestry inference (LAI) is applied to phased cohort genotypes to assign ancestry labels to chromosomal tracts. Rare variants are mapped onto inferred ancestry tracts and aggregated into ancestry-specific burden counts within user-defined genomic regions. These ancestry-resolved burdens are combined with phenotype and covariate data in association models to generate local ancestry-specific summary statistics.

To address these limitations, we developed Tractor-Burden, an ancestry-aware rare variant association framework that incorporates local ancestry directly into region-based burden testing. Our method supports flexible aggregation over any user-defined genomic region; because genes are the most widely used unit for rare variant burden analyses, we demonstrate its performance using gene-based tests throughout this study. Tractor-Burden builds upon the intuition of Tractor^22^, our local ancestry-aware GWAS framework for common variant association testing. As such, Tractor-Burden partitions gene-level burden by inferred local ancestry and jointly estimates ancestry-specific effects while adjusting for global ancestry and other covariates, improving gene discovery power under effect heterogeneity while maintaining appropriate calibration. Application to large-scale sequencing data from admixed individuals in the All of Us (AoU) Research Program^1^ demonstrates improved gene discovery and biological prioritization compared to standard methods, highlighting the importance of ancestry-aware modeling for advancing inclusive and accurate studies of complex trait genetics.

Tractor-Burden uses local ancestry calls to assign each allele within a gene to an ancestral background before aggregation (Fig. 1). For each individual and gene, qualifying rare alleles are summed separately across ancestry-specific haplotypes, producing one burden term per ancestry. These ancestry-specific burden terms are then included jointly in a regression model together with covariates. For individual *i*, the model is defined as:

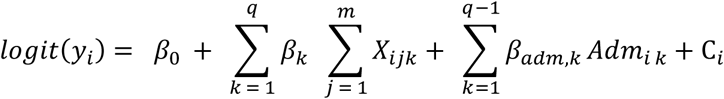

Here, *X_ijk_* represents the minor allele count for variant *j* carried on ancestry *k*, and the ancestry-specific burden effect *β_k_* captures the association between the phenotype and the aggregate burden on that ancestral background. Global ancestry terms (Adm*_ik_*) are typically included to account for phenotypic effects driven by genome-wide ancestry, while additional covariates (C*_i_*) capture non-genetic confounders. This formulation allows Tractor-Burden to estimate ancestry-specific rare-variant effects and to perform both ancestry-specific and joint gene-level tests. The framework is applicable to both binary and continuous traits.

Tractor-Burden therefore reframes rare variant aggregation in admixed cohorts from a single genome-wide burden to a set of local ancestry-resolved burdens that can be tested jointly or separately. This provides a principled framework for distinguishing shared gene-level effects from effects enriched on specific ancestral backgrounds, improving both power and interpretability when rare variant architecture differs across local ancestry tracts. Through simulations and application to whole-genome sequencing data from admixed African-European individuals in the AoU Research Program^1^, we demonstrated that Tractor-Burden maintains calibration, recovers known rare variant associations, and suggests additional gene-level signals missed by conventional burden tests. These results establish local ancestry-aware burden testing as a scalable approach for rare variant discovery in admixed genomes.

## Results

### Tractor-Burden improves power under ancestry-specific genetic architectures

To evaluate whether ancestry-aware burden modeling improves rare variant discovery in admixed populations, we performed simulations across a range of genetic architectures varying admixture proportions (Fig. 2), sample sizes (Supplementary Fig. 1), minor allele frequencies (MAF) (Fig. 2, 4), and effect sizes (Fig. 3, 4). Across demographic simulations (80/20 and 50/50 admixture scenarios), Tractor-Burden consistently improved detection power relative to ancestry-agnostic burden testing when effects differ across ancestral backgrounds, particularly when higher-impact variants are enriched within one ancestry background at higher MAF (Fig. 2). Power gains were strongest for variants with moderately rare frequencies (MAF = 0.05) compared to lower frequencies (MAF < 0.01). In the 80/20 admixture setting, models showed the greatest improvement when variants enriched in the minor ancestry drove the phenotype, reaching >80–100% power at effect sizes of ∼1–2 for MAFs ≥0.01, whereas standard burden models remained substantially underpowered. Similarly, effects restricted to the minor ancestry were best detected by LAI-tract models versus aggregate tests, although reduced ancestry proportion led to lower power. In contrast, standard burden tests diluted ancestry-specific signals across ancestral backgrounds and consistently required substantially larger effect sizes to achieve comparable power. While variants shared across ancestry groups with similar effect sizes and allele frequencies are most efficiently detected by traditional burden tests because they require fewer model parameters, Tractor-Burden exhibited only a modest reduction in power under these scenarios (Fig. 3, 4; Supplementary Fig. 3).

**Figure 2.**
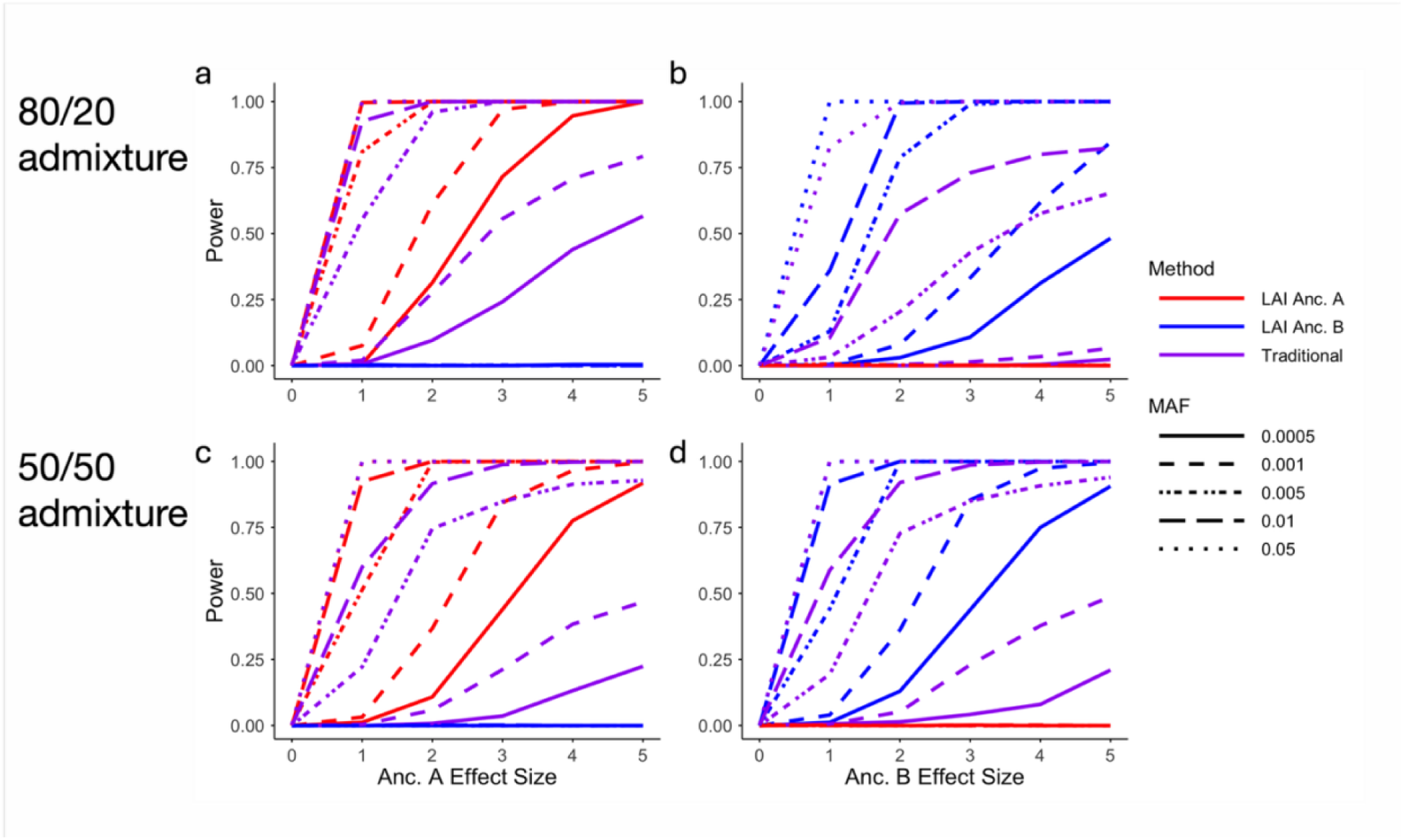
Tractor-Burden improves power for ancestry-specific rare variant association testing across admixture proportions and allele frequencies. Simulations were performed under two-way admixture models with either 80% Anc. A / 20% Anc. B admixture (**a,b**) or 50% Anc. A / 50% Anc. B admixture (**c,d**). Power was calculated as the proportion of simulations exceeding the Bonferroni-corrected significance threshold. Panels a and c show power across increasing Anc. A effect sizes while the Anc. B effect size was fixed at zero. Panels **b** and **d** show power across increasing Anc. B effect sizes while the Anc. A effect size was fixed at zero.

**Figure 3.**
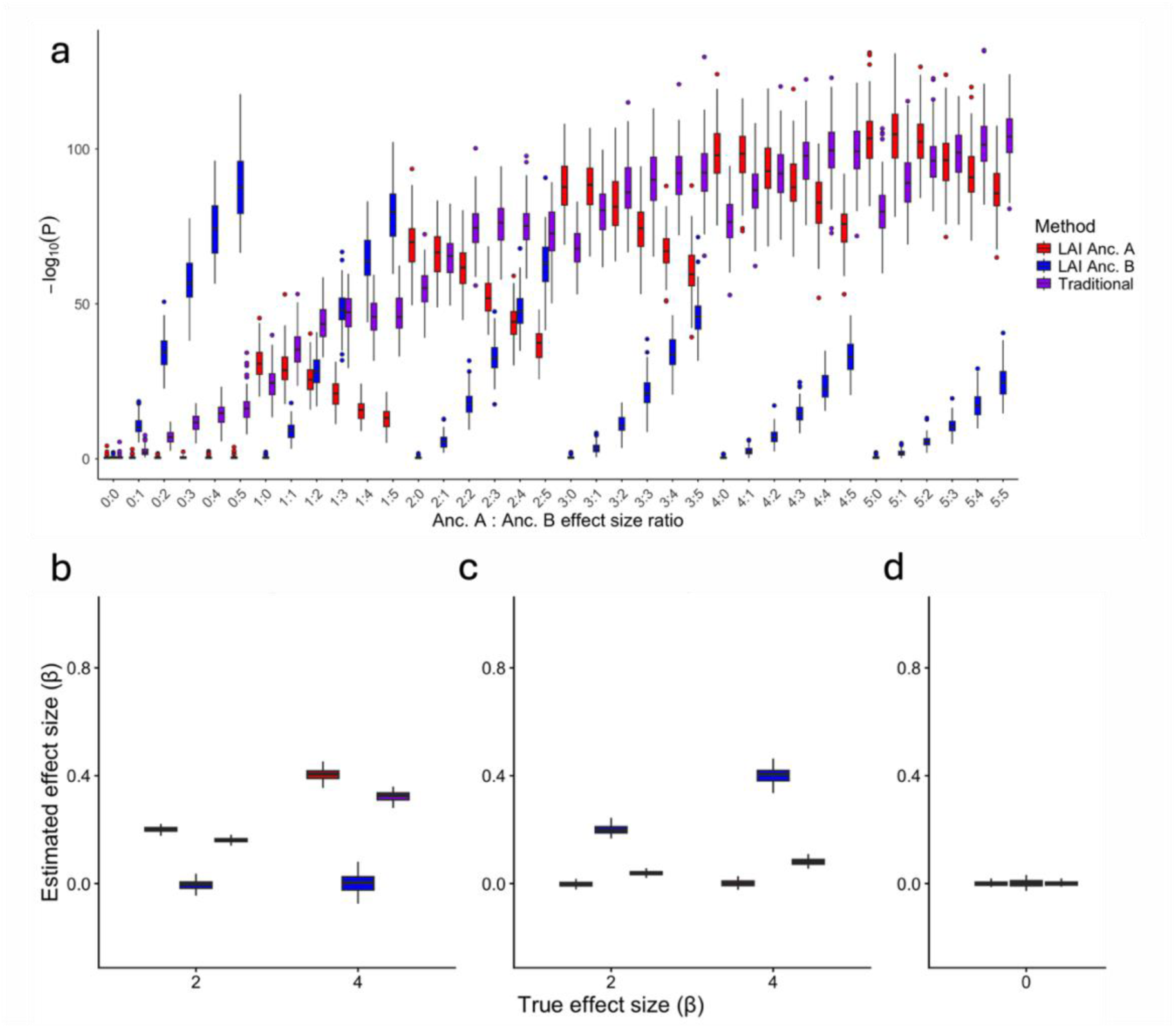
Simulation performance of Tractor-Burden under an 80% Anc. A / 20% Anc. B admixture model across varying effect size ratios. (**a**) Distribution of −log_10_(*P*)values across simulated combinations of Anc. A and Anc. B effect sizes under an 80% Anc. A / 20% Anc. B admixture model. Each x-axis category represents a unique Anc. A:Anc. B effect size combination evaluated across simulation replicates. (**b-d**) Distribution of estimated effect sizes (β̂) across simulation replicates under scenarios where causal effects were present only in Anc. A (**b**), only in Anc. B (**c**), or absent in both ancestries (**d**). Boxplots show estimated regression coefficients from the ancestry-specific and traditional burden models. The x-axis indicates the simulated true effect size (*β*), and the y-axis indicates the estimated effect size (β̂). All boxplots show the median (center line), interquartile range (box), and 1.5×interquartile range (whiskers) across 100 simulation replicates per condition, with points beyond this range plotted as outliers.

Across simulations, more common causal variants produced higher overall power to detect gene-level associations, consistent with increased variant counts across the cohort. However, the relative advantage of ancestry-aware burden modeling became increasingly apparent as variants became rarer (Fig. 2). As MAF decreased, the power difference between ancestry-specific burden models and traditional aggregate burden testing widened substantially, regardless of admixture proportions. Additional simulations varying the number of causal variants and ancestry-specific architectures demonstrated similar trends across a broad range of conditions (Supplementary Figs. 1-4). Tractor-Burden retained sensitivity even when causal variants were unevenly distributed across ancestral haplotypes.

We next evaluated Tractor-Burden across a range of ancestry-specific effect size architectures by varying the ratio of effect sizes for 80% Anc. A and 20% Anc. B admixture while holding the MAF constant (Fig. 3). When effect sizes were shared across ancestries (e.g., 1:1-5:5), traditional burden testing generally produced the strongest association signals (Fig. 3a), reflecting the increased power gained by aggregating risk alleles across ancestral backgrounds. However, as effect sizes became increasingly ancestry-specific, Tractor-Burden preferentially concentrated signal within the causal ancestry. Under Anc. A-enriched architectures (e.g., 2:0-5:0), the LAI Anc. A model achieved substantially greater significance than the LAI Anc. B model, whereas the opposite pattern was observed when effects were restricted to Anc. B (e.g., 0:2-0:5). Intermediate architectures exhibited a corresponding shift in significance toward the ancestry harboring the larger effect size. Under heterogeneous effect architectures, ancestry-specific models more accurately recovered the simulated causal effects while minimizing signal dilution across ancestral backgrounds (Fig. 3b,c). When effects were simulated exclusively on Anc. A tracts, the Anc. A-specific model recovered effect estimates close to the true simulated values (β ≈ 0.2 and 0.4), whereas the Anc. B-specific model remained centered near zero. In contrast, the ancestry-agnostic aggregate burden model produced estimates (β ≈ 0.15-0.32) consistent with dilution of ancestry-enriched effects. Similarly, under Anc. B-specific architectures, the Anc. B-specific model accurately recovered the simulated effects while Anc. A-specific estimates remained near zero, whereas aggregate burden estimates again underestimated the ancestry-specific signal. Tractor-Burden remained well calibrated under the null, with ancestry-specific and aggregate models centered near zero effect estimates in the absence of simulated signal (Fig. 3d).

To further evaluate the impact of ancestry-specific allele frequency differences on rare variant burden testing, we simulated causal variants across combinations of ancestral MAFs and numbers of causal variants (Fig. 4). For a given MAF, association significance generally increased as the number of causal variants increased from 1 to 20, reflecting the stronger aggregate burden generated by multiple risk variants. Across all simulation settings, traditional burden testing produced the most significant associations, consistent with the simulation design in which causal variants were assigned identical effect sizes across ancestries. When causal variants occurred at similar frequencies in both ancestries (Fig. 4a, e, i), the advantage of traditional burden testing was greatest because aggregating variants across ancestries maximized statistical power. As ancestry-specific MAF differences increased (Fig. 4c, f, i), the ancestry-specific Tractor-Burden model corresponding to the ancestry harboring the higher-frequency causal variants became progressively more significant. Specifically, enrichment of causal variants in Anc. B (Fig. 4b, c) preferentially strengthened the LAI Anc. B association, whereas enrichment in Anc. A (Fig. 4d and g) resulted in stronger LAI Anc. A associations. Although these ancestry-specific models remained less significant than the traditional burden test, they successfully identified the ancestral background driving the burden signal, a distinction that is not possible using conventional aggregation approaches. Thus, Tractor-Burden sacrifices only a modest amount of power to provide ancestry-specific resolution, allowing rare variant associations to be localized to the ancestral population in which causal variants are enriched.

**Figure 4.**
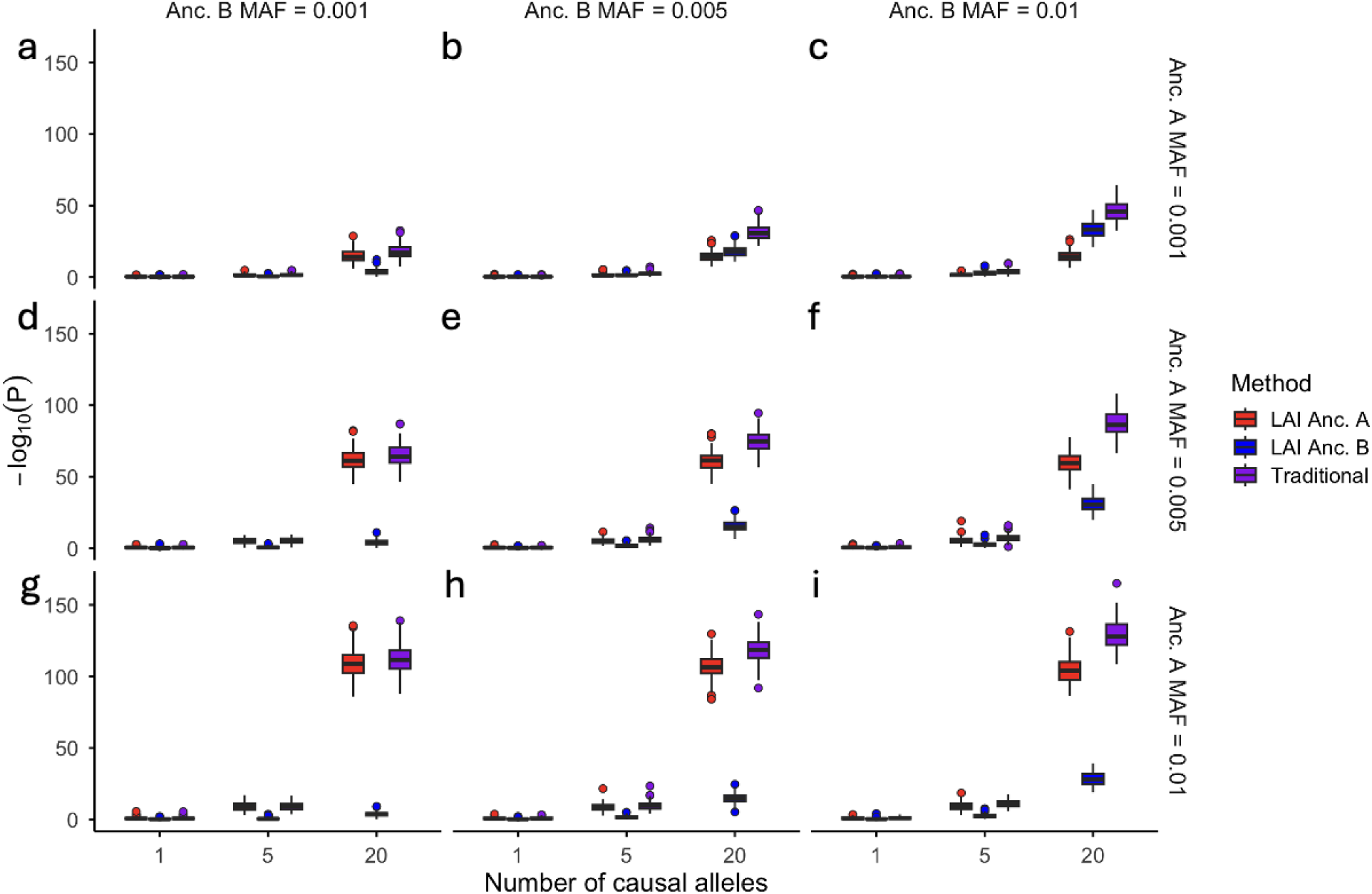
Simulation performance across varying ancestry-specific minor allele frequencies and numbers of causal alleles under an 80% Anc. A / 20% Anc. B admixture model. Simulations were performed with equal effect sizes in both ancestries (*β*_Anc.A_ = *β*_Anc.B_ = 1). Columns correspond to increasing Anc. B minor allele frequencies (MAFs) (0.001, 0.005, and 0.01), while rows correspond to increasing Anc. A MAFs (0.001, 0.005, and 0.01). Panels **a-c**, **d-f**, and **g-i** represent the combinations of ancestry-specific MAFs shown on the right and top margins. The x-axis indicates the number of simulated causal alleles (1, 5, or 20), and the y-axis shows association significance as −log_10_(*P*). All boxplots show the median (center line), interquartile range (box), and 1.5×interquartile range (whiskers) across 100 simulation replicates per condition, with points beyond this range plotted as outliers.

### Application of Tractor-Burden to admixed All of Us participants

We next applied Tractor-Burden to 47,152 African-European admixed participants from the AoU Research Program following ancestry-aware quality control and local ancestry inference procedures (Fig. 1, Supplementary Fig. 5). Rare coding variants were evaluated using ancestry-resolved burden testing and compared against conventional burden approaches including CMCWald^13^ and SKAT-O^15^ using matched variant sets and filtering criteria. Across analyses, Tractor-Burden enabled direct estimation of ancestry-specific burden effects, facilitating interpretation of whether association signals were shared across ancestral backgrounds or enriched within specific ancestry tracts.

We first evaluated low-density lipoprotein (LDL) cholesterol as a proof-of-concept phenotype, given it has a well-characterized genetic architecture across populations^23,24^. Across methods, known lipid-associated genes were recovered, including *LDLR*, providing a positive control (Fig. 5). However, Tractor-Burden further resolved these associations by partitioning signal across ancestry-specific haplotypes. For *LDLR*, the African ancestry-specific burden model demonstrated the strongest association signal across all tested methods (β = 0.411, *P* = 3.51 × 10⁻⁷; −log10(*P*) = 6.45), whereas the European ancestry-specific burden component showed no significant association (β = −0.033, *P* = 0.866). In comparison, conventional burden approaches produced weaker aggregate associations, including CMCWald^13^ (β = 0.398, *P* = 2.58 × 10⁻⁶; −log10(*P*) = 5.59) and SKAT-O^15^ (*P* = 1.04 × 10⁻⁶; −log10(*P*) = 5.98). These findings suggest that the *LDLR* association was driven primarily by rare variants residing on African ancestry haplotypes in our admixed cohort, illustrating how incorporating local ancestry information can increase statistical power for detecting known rare variant associations, even at well-established loci such as *LDLR*.

**Figure 5.**
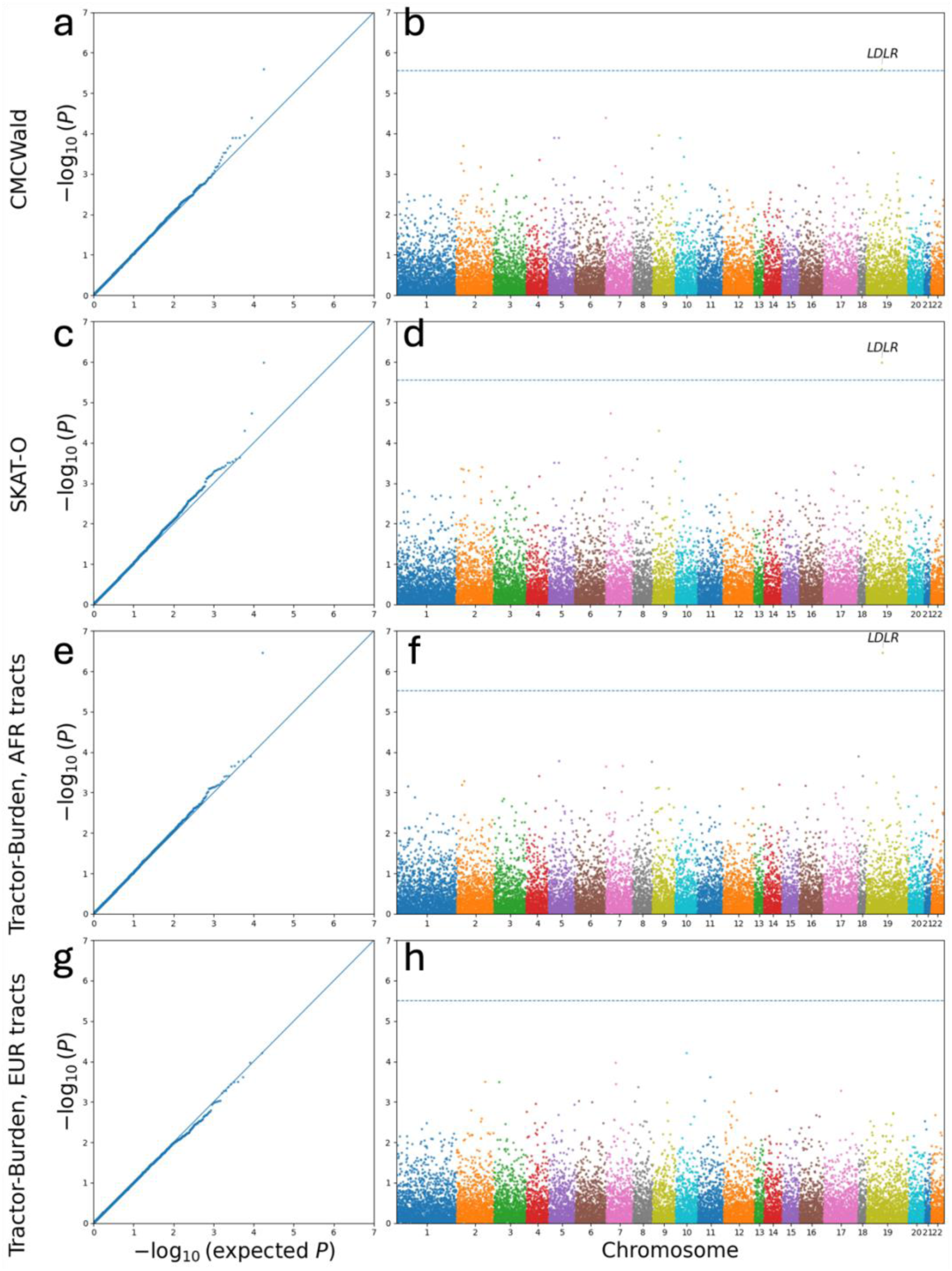
Quantile–quantile and Manhattan plots for gene-based association tests across methods for LDL cholesterol. Quantile–quantile (QQ) plots (**a,c,e,g**) and Manhattan plots (**b,d,f,h**) comparing gene-based association results across four methods: CMCWald^13^, SKAT-O^15^, Tractor-Burden in African ancestry tracts, and Tractor-Burden in European ancestry tracts. QQ plots show observed versus expected −log₁₀(P) values, with the diagonal indicating the null expectation. Manhattan plots display −log₁₀(P) values across genomic positions, with the dashed horizontal line indicating the Bonferroni significance threshold.

We next applied Tractor-Burden to type 2 diabetes (T2D) in 9,163 cases and 37,989 controls from the AoU cohort. Similar to the LDL analyses, QQ plots demonstrated appropriate calibration across methods (Supplementary Fig. 6). Ancestry-aware burden testing identified distinct association patterns relative to standard burden approaches, highlighting genes with suggestive ancestry-enriched burden signals that were less apparent in aggregate analyses. Among the suggestive genes, *USP24* demonstrated a strong ancestry-specific association within European ancestry tracts (β = 0.882, OR = 2.42, *P* = 5.0 × 10⁻⁶; −log10(*P*) = 5.34), whereas little evidence of association was observed within African ancestry tracts (β = −0.088, OR = 0.915, *P* = 0.41) (Supplementary Fig. 6). Standard burden approaches including CMCWald^13^ and SKAT-O^15^ similarly failed to detect suggestive aggregate associations for *USP24*, suggesting that ancestry-agnostic aggregation diluted the ancestry-enriched signal observed by Tractor-Burden.

Pathway enrichment analyses of AFR-specific burden signals identified significant enrichment of growth and metabolic signaling pathways, including PI3K-AKT-mTOR signaling, whereas no comparable pathway enrichment was observed using the other methods. Separately, we detected an EUR-specific signal at *USP24,* which has previously been implicated in lipid metabolism, inflammatory signaling, and insulin resistance pathways^25,26^, supporting its potential relevance to T2D-related metabolic biology. To further characterize the EUR ancestry-specific *USP24* association, we examined the individual variants contributing to the burden signal (Supplementary Table 1). The association was supported by multiple qualifying rare variants rather than a single predominant allele. Several variants were observed in multiple carriers within the All of Us cohort, whereas many additional variants were present in only one or two individuals. Comparison with UK Biobank allele frequencies showed that several contributing variants were not observed among European participants in UK Biobank, while others were present at extremely low frequencies. Together, these analyses illustrate how Tractor-Burden can distinguish shared from ancestry-enriched burden signals and prioritize genes and pathways that may be obscured by conventional aggregation.

## Discussion

RVAS have become increasingly important for understanding complex trait architecture, yet most existing analytical frameworks remain optimized for homogeneous populations. As large-scale sequencing resources and admixed populations continue to grow, there is an urgent need for methods capable of appropriately modeling the mosaic ancestry structure characteristic of admixed genomes. Here, we introduce Tractor-Burden, an ancestry-aware rare variant association framework that incorporates local ancestry directly into gene-based burden testing. Across simulations, Tractor-Burden improved statistical power under heterogeneous genetic architecture while maintaining appropriate calibration. These gains were particularly evident when allele frequencies or effect sizes differed by ancestry, conditions under which conventional burden methods frequently attenuated association signal through ancestry-agnostic aggregation. Importantly, Tractor-Burden also retained adequate discovery power when causal variants were shared across ancestries, with only a modest reduction in power. By partitioning burden signal across ancestry-specific haplotypes, Tractor-Burden enables direct estimation and interpretation of ancestry-resolved rare variant effects. This distinction is particularly relevant in admixed populations, where causal variants may differ substantially in frequency across local ancestry tracts and may thus have effects that are partially or fully ancestry-specific.

Tractor-Burden is expected to provide the greatest advantage when rare variant frequencies or effects differ across local ancestry backgrounds. In contrast, when causal variants have similar frequencies and effects across ancestries, aggregate burden tests may retain greater power because they estimate fewer parameters. Thus, Tractor-Burden is best viewed as a complementary framework that improves inference when local ancestry is informative, rather than a universal replacement for standard burden testing. Beyond power, another central advantage of Tractor-Burden is interpretability. By separating gene-level burden by local ancestry, the framework can distinguish signals that are shared across ancestry backgrounds from those driven primarily by one ancestry-specific haplotype context. This distinction can guide downstream fine-mapping, replication, and functional follow-up.

Application to whole genome sequencing data from the AoU Research Program further demonstrated the utility of ancestry-aware burden modeling at biobank scale. Tractor-Burden successfully recapitulated established associations while additionally refining ancestry-enriched burden signals that were less apparent using standard burden approaches. For example, ancestry-specific modeling localized rare variant signal at *LDLR* primarily to African ancestry tracts, whereas aggregate burden methods produced comparatively weaker associations. This observation is consistent with prior evidence demonstrating ancestry-specific differences in lipid-associated genetic architecture^27,28^. Although no significant association was detected on European local ancestry tracts, this should not be interpreted as evidence for the absence of a European effect. Because European local ancestry represents a substantially smaller proportion of chromosomes in this admixed cohort, reduced statistical power may have limited our ability to detect ancestry-specific associations on European tracts. *LDLR* is a fundamental regulator of LDL cholesterol metabolism with well-established effects across diverse ancestral groups, including large multi-ancestry sequencing and association studies that have consistently implicated both common and rare variation at this locus in lipid regulation and cardiovascular disease risk^28^. Our findings instead suggest that, within this admixed cohort, the aggregated rare variant burden contributing to the association was enriched on African ancestry tracts, illustrating how ancestry-aware burden testing can reveal ancestry-specific distributions of risk variation even within genes that are broadly relevant across populations^9^.

Similarly, our ancestry-aware analyses prioritized *USP24* as a potential ancestry-enriched T2D-associated locus within European ancestry tracts, while standard burden methods failed to detect significant aggregate associations. Variant-level examination demonstrated that this signal was supported by the combined contribution of multiple individually rare variants rather than a single dominant allele (Supplementary Table 1). Notably, several contributing variants were absent from the UK Biobank European cohort despite contributing to the burden association in All of Us. Although these observations require validation in independent datasets, they suggest that ancestry-aware analyses of admixed populations may identify rare variation that is underrepresented in homogeneous European cohorts. Importantly, this signal was identified despite European ancestry representing the minority ancestry within our study cohort, highlighting the ability of Tractor-Burden to detect ancestry-specific associations even under reduced effective sample size. In contrast, some previously reported canonical T2D burden signals, including *HNF1A*^29^, were not observed within our restricted two-way admixed cohort due to the absence of key extremely rare variants among the 47,152 individuals included in our local ancestry-informed analyses. Further investigation of the All by All T2D associations suggested that the *HNF1A* pLoF burden signal was primarily driven by the ultra-rare frameshift variant chr12-120994313-G-GC. All identified carriers of this variant within AoU belonged to predicted AMR or EUR ancestry groups, consistent with gnomAD population frequencies, and no carriers were present in our restricted AFR-EUR admixed cohort. These findings highlight how ancestry composition and ultra-rare variant representation can influence rare variant burden signal detection across studies.

Several limitations and practical considerations should be considered when applying Tractor-Burden across diverse cohorts and study designs. First, Tractor-Burden depends on accurate local ancestry inference, and performance may be sensitive to reference panel quality, phasing accuracy, and local ancestry uncertainty^30^. Because rare variants are projected onto inferred ancestry tracts, inaccuracies in LAI may propagate into downstream burden estimates, particularly in genomic regions with sparse informative markers, potentially reducing discovery power^22^. To promote reliable ancestry-specific burden estimation, we recommend restricting analyses to ancestries that comprise at least 5% of the study population and requiring individuals to have at least 5% representation from each modeled ancestry. Second, we primarily evaluated two-way African-European admixture; additional benchmarking will be necessary to assess performance in more complex multi-way admixed populations. Third, ancestry-specific burden estimation may become unstable when variant counts are extremely low or ancestry components are poorly represented. Accordingly, carrier counts within each ancestry-specific burden should be evaluated and ancestry-specific effect estimates interpreted cautiously when one burden component is sparse. Appropriate variant selection is also important. MAF thresholds should be tailored to the phenotype and genetic architecture under study, as overly permissive thresholds may dilute burden signals. Likewise, variant annotation filters should reflect the analysis objective; restricting analyses to higher-confidence functional consequences, such as SnpEff^31^ high- and moderate-impact variants, may improve power and interpretability in some settings. Although we focused on gene-based burden masks defined by rare coding variant annotations, Tractor-Burden is agnostic to both the choice of variants and the genomic regions being tested, and can readily incorporate alternative variant filtering strategies, weighting schemes, functional annotations, and user-defined genomic intervals. Finally, future work could extend Tractor-Burden to mixed-model or saddlepoint-based frameworks to better accommodate relatedness, case-control imbalance, and extremely low carrier counts. To facilitate reproducible application, Tractor-Burden is accompanied by detailed documentation, a tutorial, and a toy dataset freely available through GitHub (https://github.com/Atkinson-Lab/Tractor-Burden).

As genomic resources continue to expand globally, methods capable of modeling ancestry-resolved genetic architecture will be essential for equitable genomic discovery. Tractor-Burden provides a scalable framework for ancestry-aware rare variant association testing and supports more inclusive characterization of complex trait genetics across populations. More broadly, we anticipate that local ancestry-aware rare variant analyses will improve our understanding of when gene-level effects are shared across populations versus enriched on specific ancestry backgrounds, clarify the role of local ancestry in rare variant association signals, and support interpretation of rare variant findings across cohorts with different ancestry compositions. By resolving ancestry-specific burden signals that may be diluted by conventional aggregation, Tractor-Burden extends rare variant discovery to admixed genomes and provides a framework for more precise genetic analysis in biobank-scale sequencing studies.

## Methods

### Ethics statement

This study was conducted using de-identified participant data accessed through the All of Us Research Program Researcher Workbench. All participants provided informed consent for participation in the All of Us Research Program. The Baylor College of Medicine Institutional Review Board determined that this project did not constitute human subjects research, and no additional IRB approval was required.

### Simulations for Tractor-Burden

We conducted simulation studies to evaluate the performance of Tractor-Burden under a range of genetic architectures representative of admixed populations. Simulations were performed using simulated two-way admixed cohorts with ‘A’ and ‘B’ ancestry components, modeling both 80:20 and 50:50 admixture proportions. Individual-level ancestry proportions were generated using a beta distribution, and local ancestry at each locus was simulated using a binomial distribution (n = 2) to assign ancestry-specific haplotypes.

We simulated gene regions containing multiple variants, with each variant independently assigned ancestry-specific minor allele frequencies (MAF) spanning 0.0005 to 0.05. Rare variants were defined across this spectrum to reflect realistic allele frequency distributions observed in sequencing studies. For each simulation, a subset of variants (n = 1, 5, or 20) was designated as causal. Effect sizes (β) were varied across a range from 0 to 5, and ancestry-specific effect configurations were explored, including scenarios where effects were present in only one ancestry or differed between ancestries. Additional simulations included matched effect sizes across ancestries with varying MAF, and matched MAF with varying effect sizes. Phenotypes were simulated for continuous traits, generated as a function of ancestry-specific genetic burden and global ancestry proportion. All simulations were conducted at a fixed sample size of N = 10,000.

To evaluate performance across various realistic scenarios, we conducted 100 independent simulation replicates for each parameter combination. Parameter grids included variation in ancestry proportions (80:20 and 50:50), MAF distributions, effect sizes, and the number of causal variants. Simulations were designed to maintain a false discovery rate consistent with field standards. All simulations were performed in R (version 2025.9.0.387)^33^.

### Whole-genome sequencing data processing and quality control

Whole-genome sequencing (WGS) data were obtained from the All of Us (AoU) Research Program^1^ (Controlled Tier, version 8; GRCh38-aligned). Analyses were restricted to unrelated individuals with high-quality genotype data and available demographic information. Variant-level quality control included restriction to biallelic SNPs passing site- and genotype-level filters, with variant missingness <1% and sample missingness <1%. For common variant analyses used in downstream ancestry inference, variants were further filtered to MAF >0.005. A reference panel was constructed from high-coverage (30×) 1000 Genomes Project samples aligned to GRCh38 and phased by the International Genome Sample Resource ^34^. Autosomal SNPs with MAF >0.05 were linkage disequilibrium (LD)-pruned (500 kb window, r² < 0.1), excluding long-range LD regions, and principal components (PCs) were computed for ancestry inference.

### Selection of AFR-EUR two-way admixed individuals

We analyzed a subset of 47,152 two-way African-European (AFR-EUR) admixed individuals from the AoU Research Program^1^ defined using previously established ancestry and sample quality control procedures. Briefly, analyses were restricted to unrelated individuals passing standard data quality filters and with available sex-at-birth information.

Genetic ancestry was inferred using principal component analysis (PCA) based on a reference panel derived from 1000 Genomes Project^34^ populations. AFR and EUR reference individuals included populations from West African (ESN, GWD, MSL, YRI) and European (CEU, GBR, IBS, TSI) groups. Using the first four principal components, study participants were projected onto the axis defined by the mean positions of AFR and EUR reference samples using the admix-kit framework. The position along this axis reflects relative AFR–EUR ancestry proportions, while the orthogonal distance quantifies deviation from the two-way admixture cline.

We retained individuals consistent with two-way AFR–EUR admixture by requiring at least 5% AFR ancestry and excluding individuals with substantial contributions (>5%) from other ancestral backgrounds. Individuals with large deviations from the AFR–EUR cline (normalized distance >1.5) were excluded to ensure alignment with the reference populations.

### Local ancestry inference using a curated reference panel

Local ancestry inference (LAI) was performed on the AoU ^1^ cohort using a previously established pipeline based on phased genotype data and AFR and EUR reference panels derived from the 1000 Genomes Project^34^. Briefly, SNPs were harmonized between datasets and LAI was conducted using FLARE^35^. Inferred ancestry proportions were validated against global ancestry estimates obtained using ADMIXTURE^36^ and Rye, with high concordance across methods. To reduce edge effects and instability in low-quality regions, the first and last 1 Mb of each chromosome were excluded.

### Local ancestry assignment for rare coding variants

Rare variant analyses focused on coding variants within CCDS exonic regions of the 47,152 two-way AFR–EUR admixed individuals. Variants were filtered to exclude common variation, defined as population-specific allele frequency >1% or allele count >100 in the ACAF callset. Local ancestry assignments derived from common variants were projected onto rare variants using a nearest-marker approach, assigning ancestry labels based on flanking inferred tracts. Using these assignments, ancestry-specific variant representations were generated with Tractor^22^, producing ancestry-resolved dosage and haplotype count files for downstream gene-based association analyses.

### Phenotype definition and cohort selection

Phenotypes were derived from AoU electronic health records and laboratory measurements. For LDL cholesterol (N = 10,855), measurements were defined using the OMOP concept “Cholesterol in LDL (Low-Density Lipoprotein) [Mass/volume] in Serum or Plasma” (measurement_concept_id = 3028288) with units restricted to milligrams per deciliter (unit_concept_id = 8840). Extreme values beyond ±3 standard deviations were excluded, followed by inverse rank-normal transformation. For type 2 diabetes (T2D; N = 9,163 cases, 37,989 controls), case-control status was defined based on clinical diagnoses using the SNOMED concept “Type 2 diabetes mellitus” (concept_id = 201826; concept_code = 44054006). Analyses were restricted to individuals with complete phenotype and covariate data, including age at data collection and sex at birth.

### Burden Testing Parameters in All of Us Research Program

Tractor-Burden was applied to perform gene-based association testing using ancestry-specific rare variant burden models. Variants were annotated using SnpEff^31^, and analyses were restricted to high- and moderate-impact functional annotations. For each gene, ancestry-specific burden scores were constructed and modeled jointly within a regression framework, allowing estimation of ancestry-specific effect sizes. Association testing included covariates for age, sex, and global African ancestry proportion. This framework enables detection of ancestry-enriched rare variant effects while accounting for heterogeneity in allele frequency and effect size across ancestral backgrounds.

For comparison, we benchmarked Tractor-Burden against two established rare variant association approaches implemented in RVTESTS^37^: CMCWald^13^ and SKAT-O^15^. Both methods were run on the same filtered variant sets used in Tractor-Burden to enable a direct comparison of association performance. Analyses were restricted to rare coding variants passing our ACAF filtering strategy and annotated with SnpEff^31^ “High” or “Moderate” predicted functional consequences. Variants were grouped at the gene level using refFlat-based gene definitions, and only variants with minor allele count (MAC) ≥1 were retained (--siteMACMin 1). Association models included the same phenotype and covariate framework as Tractor-Burden, adjusting for age, sex, and global African ancestry proportion within the AFR–EUR admixed cohort.

Statistical significance was determined using a Bonferroni correction based on the number of genes evaluated in each analysis. Bonferroni-adjusted significance thresholds are indicated in all Manhattan plots. For T2D analyses, genes exceeding a suggestive significance threshold of −log10(P) ≥ 3.25 were additionally highlighted to facilitate interpretation of sub-threshold association signals.

## Data availability

This study used data from the All of Us Research Program’s Controlled Tier Dataset version 8 (GRCh38-aligned) available to authorized users on the Researcher Workbench. Summary statistics are freely available at https://github.com/Atkinson-Lab/Tractor-Burden-Simulations-Summary-Stats.

## Code availability

Code is freely available at https://github.com/Atkinson-Lab/Tractor-Burden.

## Acknowledgments

We gratefully acknowledge All of Us participants for their contributions, without whom this research would not have been possible. We also thank the National Institutes of Health’s All of Us Research Program for making the cohort data examined in this study available. This work was funded by the National Institutes of Health (K01 MH121659 and R01 HG012869 to E.G.A.; P.K. was supported by T32 GM139534-03 and T32 GM136554-05).

## Author Contributions

Conceptualization/Funding acquisition/Supervision: E.G.A. Data curation/software/investigation: P.K. Formal analysis P.K. and T.T. Validation: P.K. and A.M. Methodology: P.K., T.T., and E.G.A. Visualization: P.K. and W.L. Writing-original draft: P.K. Writing-review and editing: P.K., T.T., W.L., A.M., L.H., N.C., W.Z., R.S.D., and E.G.A.

