## Supplementary Information for "Local ancestry-informed rare variant burden testing improves gene discovery in admixed populations"

**SUPPLEMENTARY TABLE**

**Supplementary Table 1. Variant-level characterization of the EUR ancestry-specific USP24 burden signal associated with type 2 diabetes in the All of Us Research Program.** Variant-level summary of rare USP24 variants contributing to the EUR ancestry-specific burden association with type 2 diabetes identified by Tractor-Burden. All contributing variants were annotated as missense variants. For each qualifying variant, the table reports the local ancestry tract on which the variant was observed, the number of carriers within the All of Us cohort, case and control carrier counts, and corresponding allele count and minor allele frequency in the UK Biobank European (EUR) and African (AFR) populations. Variants highlighted in gray were not observed in the UK Biobank European dataset, despite contributing to the EUR ancestry-specific burden signal detected in the admixed All of Us cohort.

| Gene | Variant | Local ancestry tract | AoU tract carriers | AoU case carriers | AoU control carriers | UKBB EUR Allele count | UKBB EUR MAF | UKBB AFR Allele count | UKBB AFR MAF | Interpretation |
|---|---|---|---:|---:|---:|---|---|---|---|---|
| USP24 | chr1:55124498:G:A | EUR | 7 | 3 | 4 | 179 | 0.0002 | 0 | 0 | Slight control enrichment among EUR carriers |
| USP24 | chr1:55214845:C:T | EUR | 7 | 6 | 1 | 77 | 0.000084 | 0 | 0 | Strong case enrichment among EUR carriers |
| USP24 | chr1:55069095:G:A | EUR | 6 | 2 | 4 | 70 | 0.000076 | 0 | 0 | Modest carrier count |
| USP24 | chr1:55121461:A:C | EUR | 4 | 0 | 4 | 72 | 0.000078 | 0 | 0 | Present only in controls |
| USP24 | chr1:55083310:T:C | EUR | 4 | 1 | 3 | 8 | 0.000008 | 0 | 0 | Modest carrier count |
| USP24 | chr1:55154178:C:T | EUR | 4 | 2 | 2 | 90 | 0.000098 | 0 | 0 | Even distribution |

| | | | | | | | | | | |
|---|---|---|---|---|---|---|---|---|---|---|
| | | | | | | | | | | between cases and controls |
| USP24 | chr1:55172453:A:G | EUR | 4 | 2 | 2 | 139 | 0.00015 | 0 | 0 | Even distribution between cases and controls |
| USP24 | chr1:55095286:C:T | EUR | 4 | 1 | 3 | 254 | 0.00027 | 1 | 0.00005 | Predominantly observed in controls |
| USP24 | chr1:55141719:C:T | EUR | 3 | 0 | 3 | 6 | 0.000006 | 0 | 0 | Control-only carrier |
| USP24 | chr1:55094082:G:C | EUR | 3 | 1 | 2 | 10 | 0.00001 | 0 | 0 | Slight control enrichment |
| USP24 | chr1:55107326:T:C | EUR | 3 | 2 | 1 | 10 | 0.00001 | 0 | 0 | Mild case enrichment |
| USP24 | chr1:55096579:T:C | EUR | 3 | 1 | 2 | 4 | 0.000004 | 0 | 0 | Slight control enrichment |
| USP24 | chr1:55124526:T:C | EUR | 3 | 2 | 1 | 31 | 0.00003 | 0 | 0 | Mild case enrichment |
| USP24 | chr1:55129537:T:C | EUR | 2 | 2 | 0 | 9 | 0.000009 | 0 | 0 | Observed only in cases (very small carrier count) |
| USP24 | chr1:55132591:G:C | EUR | 2 | 1 | 1 | 15 | 0.00001 | 0 | 0 | Balanced distribution |
| USP24 | chr1:55075508:T:C | EUR | 2 | 0 | 2 | 6 | 0.000006 | 0 | 0 | Control-only carrier |
| USP24 | chr1:55159642:C:T | EUR | 2 | 0 | 2 | 69 | 0.00007 | 0 | 0 | Control-only carrier |
| USP24 | chr1:55137570:G:A | EUR | 2 | 0 | 2 | 4 | 0.000004 | 0 | 0 | Control-only carrier |
| USP24 | chr1:55129505:C:T | EUR | 2 | 1 | 1 | 26 | 0.000028 | 0 | 0 | Balanced distribution |
| USP24 | chr1:55103977:C:T | EUR | 2 | 0 | 2 | 7 | 0.000007 | 0 | 0 | Control-only carrier |
| USP24 | chr1:55125499:T:C | EUR | 2 | 0 | 2 | 116 | 0.00012 | 0 | 0 | Control-only carrier |
| USP24 | chr1:55154187:C:G | EUR | 2 | 1 | 1 | 0 | 0 | 0 | 0 | NA |

| | | | | | | | | | | |
|---|---|---|---|---|---|---|---|---|---|---|
| USP24 | chr1:55215091:T:C | EUR | 2 | 0 | 2 | 292 | 0.0003 | 1 | 0.00005 | Control-only carrier |
| USP24 | chr1:55098037:G:C | EUR | 2 | 0 | 2 | 111 | 0.0001 | 2 | 0.0001 | Control-only carrier |
| USP24 | chr1:55138955:T:C | EUR | 1 | 0 | 1 | 14 | 0.000015 | 0 | 0 | Control-only carrier |
| USP24 | chr1:55083788:T:A | EUR | 1 | 0 | 1 | 3 | 0.000003 | 0 | 0 | Control-only carrier |
| USP24 | chr1:55134164:C:T | EUR | 1 | 1 | 0 | 1 | 0.000001 | 0 | 0 | Case-only carrier |
| USP24 | chr1:55098028:C:G | EUR | 1 | 1 | 0 | 1 | 0.000001 | 0 | 0 | Case-only carrier |
| USP24 | chr1:55097066:C:T | EUR | 1 | 0 | 1 | 3 | 0.000003 | 0 | 0 | Control-only carrier |
| USP24 | chr1:55177981:T:C | EUR | 1 | 0 | 1 | 42 | 0.00004 | 0 | 0 | Control-only carrier |
| USP24 | chr1:55147708:C:A | EUR | 1 | 1 | 0 | 0 | 0 | 0 | 0 | Other pop group had an AC |
| USP24 | chr1:55095333:A:G | EUR | 1 | 0 | 1 | 10 | 0.00001 | 0 | 0 | Control-only carrier |
| USP24 | chr1:55081396:C:T | EUR | 1 | 1 | 0 | 8 | 0.000008 | 0 | 0 | Case-only carrier |
| USP24 | chr1:55141670:A:G | EUR | 1 | 1 | 0 | 10 | 0.00001 | 0 | 0 | Case-only carrier |
| USP24 | chr1:55159024:T:G | EUR | 1 | 0 | 1 | 0 | 0 | 0 | 0 | NA |
| USP24 | chr1:55159003:C:T | EUR | 1 | 0 | 1 | 6 | 0.0000065 | 1 | 0.000054 | Control-only carrier |
| USP24 | chr1:55158990:C:T | EUR | 1 | 0 | 1 | 4 | 0.0000043 | 0 | 0 | Control-only carrier |
| USP24 | chr1:55147735:C:T | EUR | 1 | 1 | 0 | 3 | 0.000003 | 0 | 0 | Case-only carrier |
| USP24 | chr1:55176392:G:A | EUR | 1 | 1 | 0 | 0 | 0 | 6 | 0.0003 | Case-only carrier |
| USP24 | chr1:55171615:C:A | EUR | 1 | 0 | 1 | 0 | 0 | 0 | 0 | NA |
| USP24 | chr1:55134092:T:C | EUR | 1 | 0 | 1 | 1 | 0.000001 | 0 | 0 | Control-only carrier |

| | | | | | | | | | | |
|---|---|---|---|---|---|---|---|---|---|---|
| USP24 | chr1:55132561:A:G | EUR | 1 | 1 | 0 | 18 | 0.000019 | 0 | 0 | Case-only carrier |
| USP24 | chr1:55215090:G:T | EUR | 1 | 0 | 1 | 0 | 0 | 2 | 0.0001 | Control-only carrier |
| USP24 | chr1:55125696:T:C | EUR | 1 | 1 | 0 | 1 | 0.000001 | 0 | 0 | Case-only carrier |
| USP24 | chr1:55129500:G:T | EUR | 1 | 0 | 1 | 2 | 0.000002 | 0 | 0 | Control-only carrier |
| USP24 | chr1:55214804:C:T | EUR | 1 | 0 | 1 | 1 | 0.000001 | 0 | 0 | Control-only carrier |
| USP24 | chr1:55120618:T:C | EUR | 1 | 1 | 0 | 2 | 0.000002 | 1 | 0.00005 | Case-only carrier |
| USP24 | chr1:55138640:A:G | EUR | 1 | 0 | 1 | 0 | 0 | 0 | 0 | NA |
| USP24 | chr1:55129550:T:C | EUR | 1 | 0 | 1 | 10 | 0.00001 | 0 | 0 | Control-only carrier |
| USP24 | chr1:55138633:G:A | EUR | 1 | 0 | 1 | 19 | 0.00002 | 0 | 0 | Control-only carrier |
| USP24 | chr1:55144146:T:C | EUR | 1 | 0 | 1 | 1 | 0.000001 | 0 | 0 | Control-only carrier |
| USP24 | chr1:55214986:T:C | EUR | 1 | 0 | 1 | 18 | 0.000019 | 0 | 0 | Control-only carrier |
| USP24 | chr1:55132558:A:C | EUR | 1 | 0 | 1 | 27 | 0.000029 | 0 | 0 | Control-only carrier |
| USP24 | chr1:55214972:G:C | EUR | 1 | 1 | 0 | 0 | 0 | 0 | 0 | NA |
| USP24 | chr1:55172409:T:C | EUR | 1 | 0 | 1 | 12 | 0.000013 | 0 | 0 | Control-only carrier |
| USP24 | chr1:55171591:T:C | EUR | 1 | 0 | 1 | 0 | 0 | 0 | 0 | NA |
| USP24 | chr1:55154244:T:C | EUR | 1 | 0 | 1 | 8 | 0.000008 | 0 | 0 | Control-only carrier |
| USP24 | chr1:55171590:A:G | EUR | 1 | 0 | 1 | 23 | 0.00002 | 0 | 0 | Control-only carrier |
| USP24 | chr1:55125481:C:T | EUR | 1 | 1 | 0 | 0 | 0 | 0 | 0 | NA |
| USP24 | chr1:55147725:C:T | EUR | 1 | 0 | 1 | 0 | 0 | 0 | 0 | Other pop group had an AC |
| USP24 | chr1:55107394:G:A | EUR | 1 | 0 | 1 | 13 | 0.000014 | 0 | 0 | Control-only carrier |

| | | | | | | | | | | |
|---|---|---|---|---|---|---|---|---|---|---|
| USP24 | chr1:55214915:C:T | EUR | 1 | 1 | 0 | 2 | 0.0000021 | 0 | 0 | Case-only carrier |
| USP24 | chr1:55125447:C:T | EUR | 1 | 0 | 1 | 31 | 0.000033 | 0 | 0 | Control-only carrier |

**SUPPLEMENTARY FIGURES**

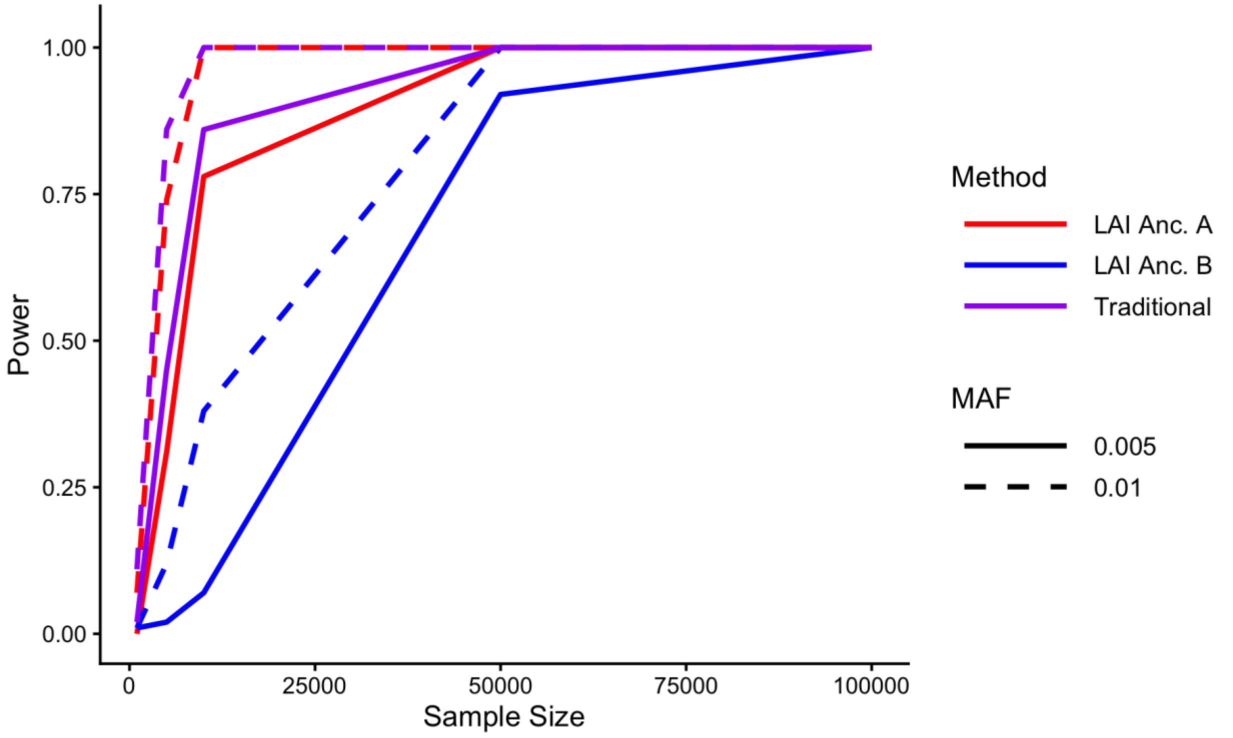

**Supplementary Figure 1: Tractor-Burden maintains high power across sample sizes, with reduced power in the underrepresented ancestry.** Power to detect rare variant burden associations in an 80:20 admixed population as a function of sample size. Red and blue lines represent ancestry-specific Tractor-Burden tests for Anc. A and Anc. B, respectively, and purple lines represent traditional burden testing. Solid and dashed lines correspond to causal variant MAFs of 0.005 and 0.01, respectively.

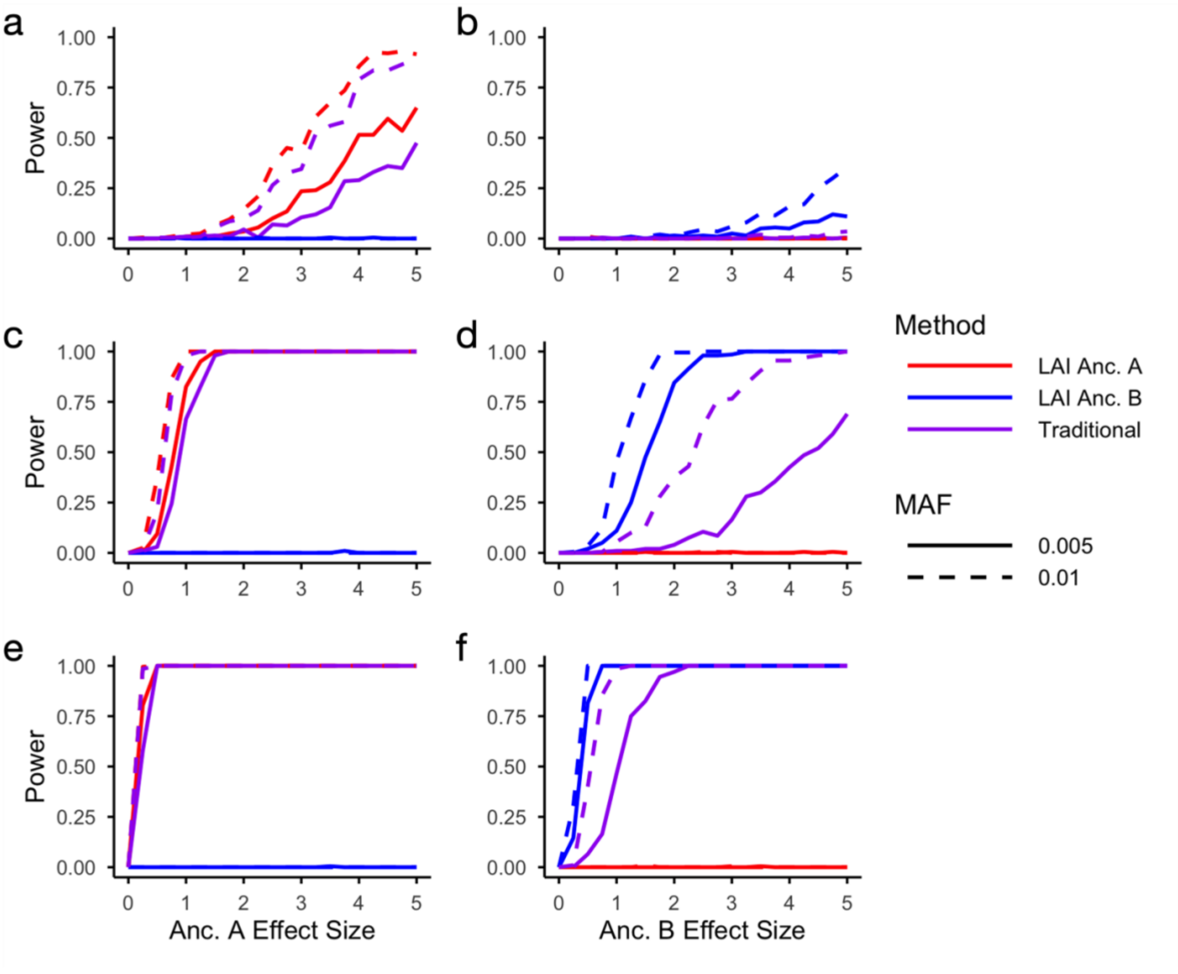

**Supplementary Figure 2: Tractor-Burden models maximize power for ancestry-enriched effects in an 80:20 admixed population.** Power as a function of simulated ancestry-specific effect size. Red, blue, and purple lines denote Anc. A-specific, Anc. B-specific, and traditional burden tests, respectively. Solid and dashed lines correspond to causal variant MAFs of 0.005 and 0.01. Panels a-b show simulations with 1 causal variant, panels c-d with 5 causal variants, and panels e-f with 20 causal variants. In panels a, c, and e, effect sizes were varied on Anc. A tracts, whereas in panels b, d, and f, effect sizes were varied on Anc. B tracts. Power increased with effect size, MAF, and the number of causal variants, with the ancestry-specific Tractor-Burden model achieving the greatest power when effects were confined to its corresponding ancestry.

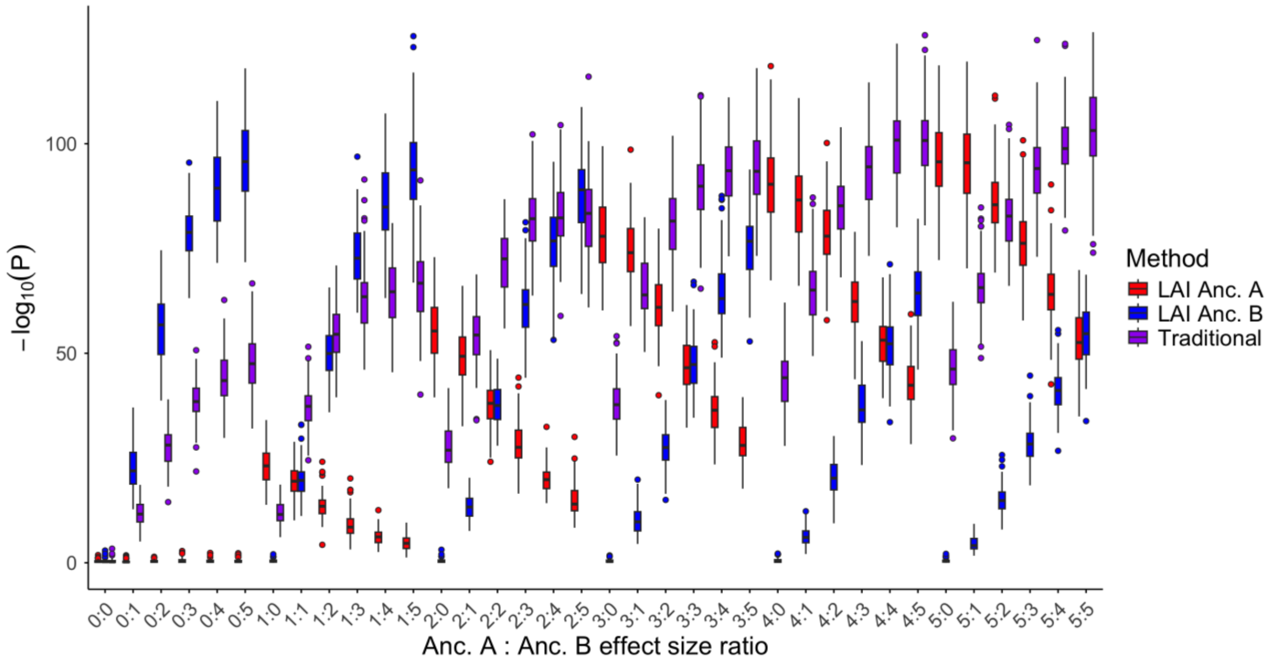

**Supplementary Figure 3: Tractor-Burden localizes ancestry-specific burden signals in a balanced 50:50 admixed population.** Association significance ($-\log_{10}P$) across simulated Anc. A:Anc. B effect size ratios in a 50:50 admixed population. Red and blue boxplots represent ancestry-specific Tractor-Burden tests for Anc. A and Anc. B, respectively, while purple boxplots represent traditional burden testing. As effect sizes became increasingly enriched in one ancestry, the corresponding ancestry-specific Tractor-Burden model achieved greater significance, whereas traditional burden testing remained most powerful when effects were shared across ancestries.

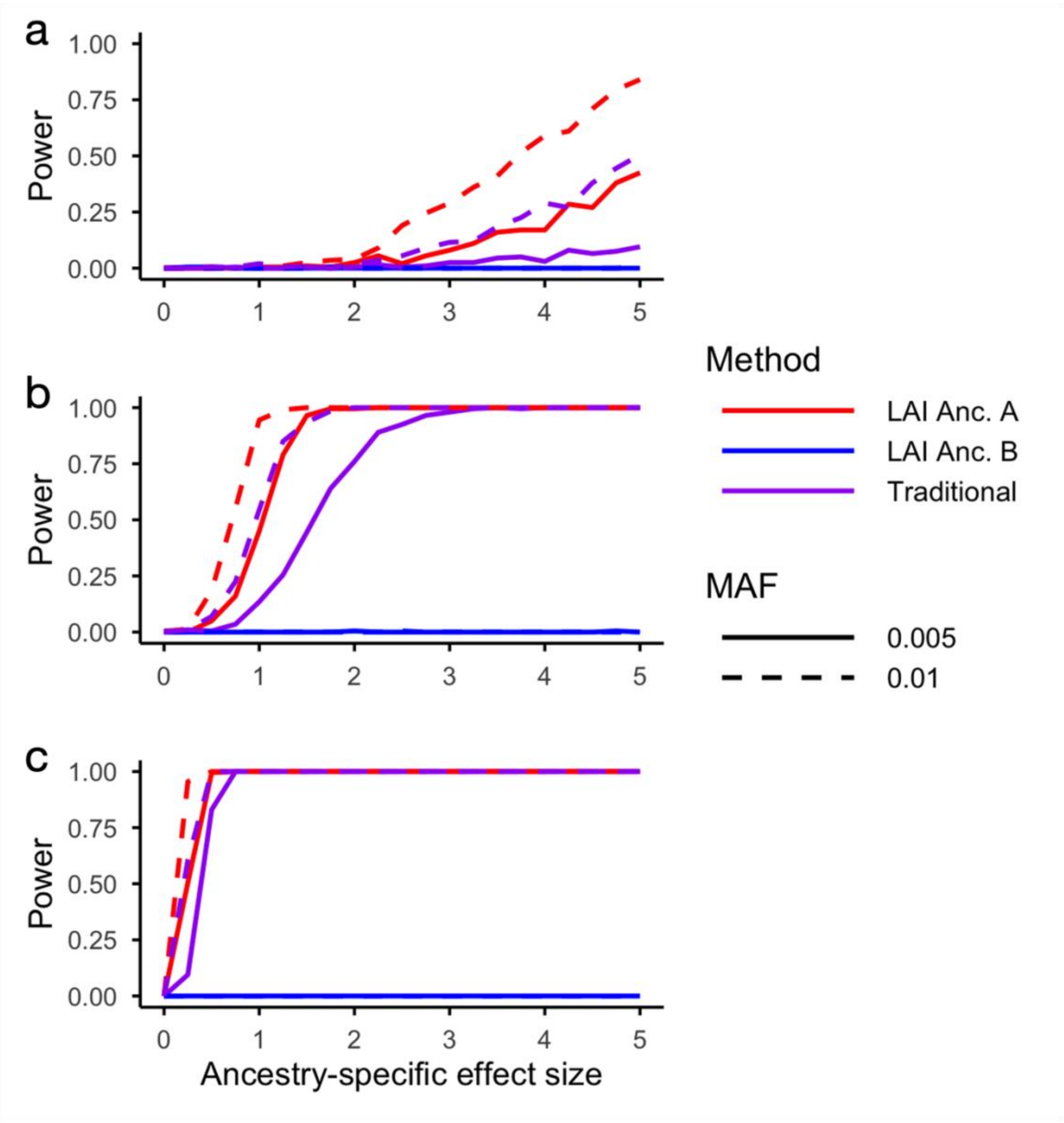

**Supplementary Figure 4: Power increases with effect size and causal variant burden for ancestry-specific Tractor-Burden tests.** Power as a function of simulated Anc. A-specific effect size (Anc. B effect size = 0) in a 50:50 admixed population. Red, blue, and purple lines denote ancestry-specific Tractor-Burden tests for Anc. A and Anc. B, respectively, and traditional burden testing. Solid and dashed lines correspond to causal variant MAFs of 0.005 and 0.01. Panels a-c represent simulations with 1, 5, and 20 causal variants, respectively.

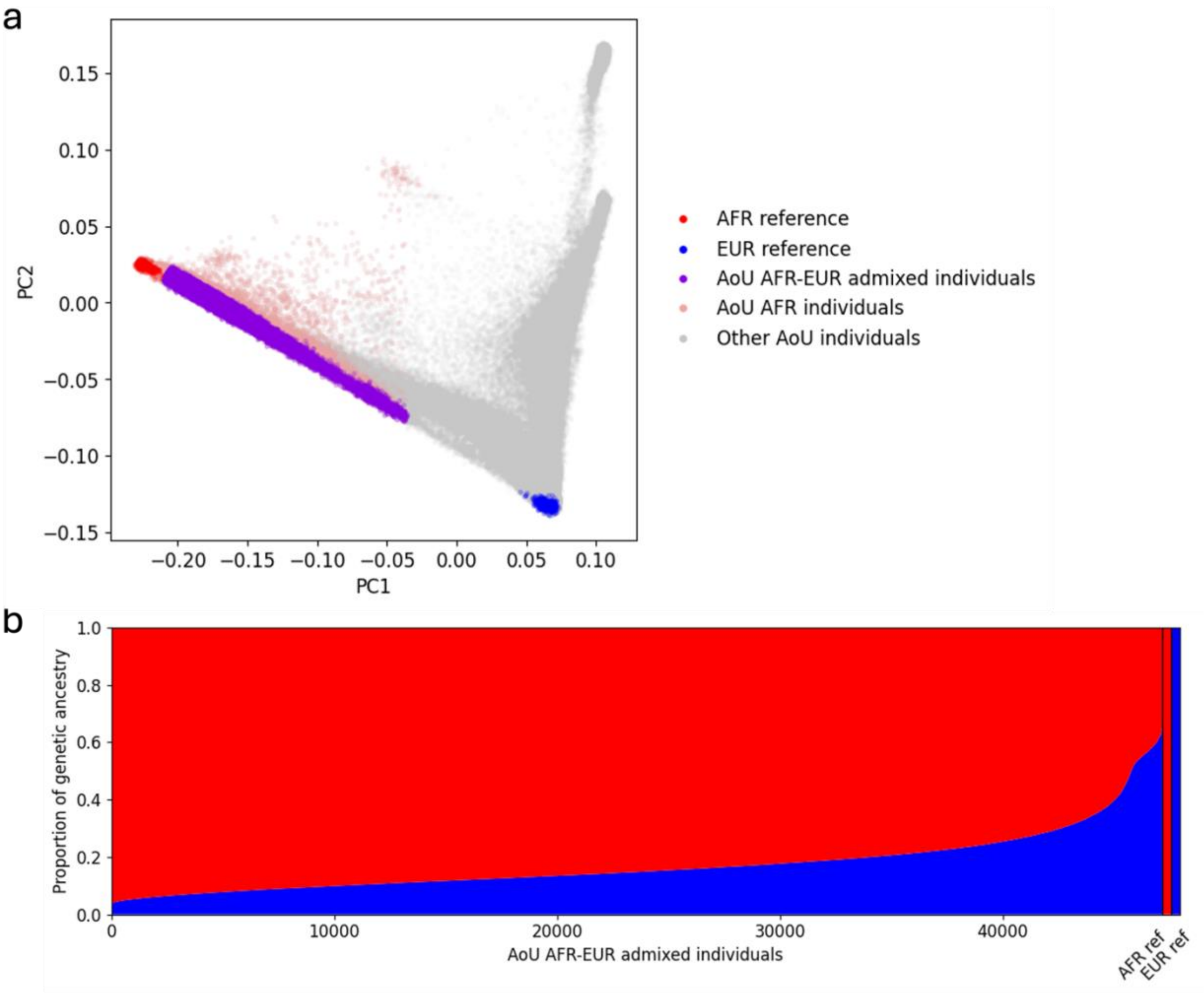

**Supplementary Figure 5: Principal component analysis and global ancestry composition of the AFR–EUR admixed All of Us [1] cohort.** Principal component analysis (**a**) of All of Us participants projected onto AFR and EUR reference populations. Purple points represent individuals selected for the AFR–EUR admixed analysis cohort, positioned along the continuum between AFR (red) and EUR (blue) reference samples. Pink points denote non-admixed AFR individuals, and gray points represent all other All of Us participants. Global ancestry estimates for the AFR–EUR admixed cohort are shown (**b**) with each vertical bar representing one individual.

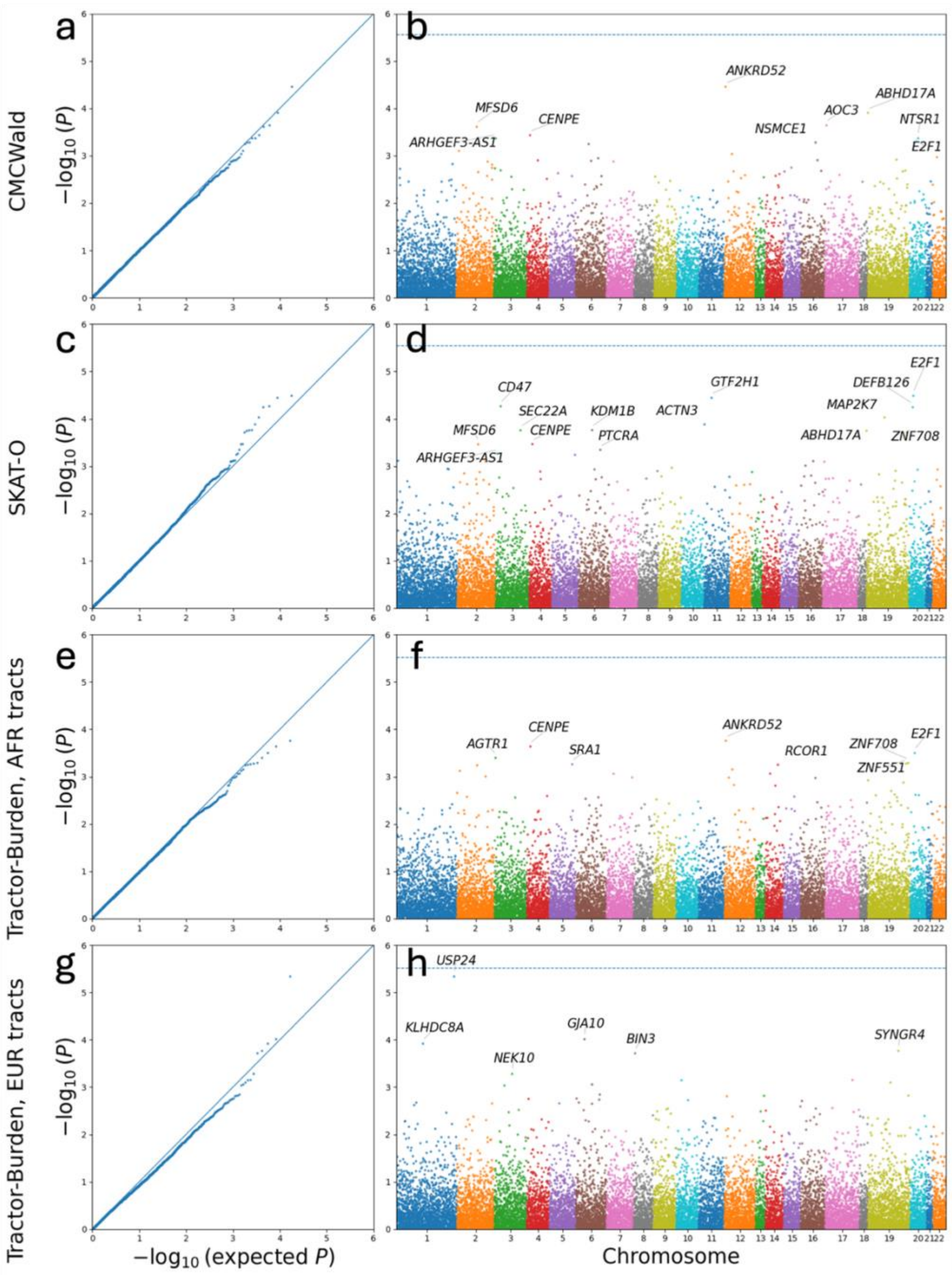

**Supplementary Figure 6. Quantile–quantile and Manhattan plots for gene-based association tests across methods for type 2 diabetes.** Quantile–quantile (QQ) plots (**a,c,e,g**) and Manhattan plots (**b,d,f,h**) comparing gene-based association results across four methods: CMCWald[13], SKAT-O[15], Tractor-Burden in African ancestry tracts, and Tractor-Burden in European ancestry tracts. QQ plots show observed versus expected $-\log_{10}(P)$ values, with the diagonal indicating the null expectation. Manhattan plots display $-\log_{10}(P)$ values across genomic positions, with the dashed horizontal line indicating the Bonferroni significance threshold.
